# Speaking the Language of Inclusion: Examining English Languages Requirements in Cardiovascular Digital Health Trials

**DOI:** 10.1101/2025.06.10.25329222

**Authors:** Narayan Schütz, Ritu Doijad, Ahmed A. Eltahir, Ashish Sarraju, Daniel Seung Kim, Fatima Rodriguez

## Abstract

**Background:** Cardiovascular medicine is rapidly evolving, as it integrates digital technologies intended to decentralize care from the clinic and/or hospital setting into patient’s homes and communities. However, clinical trials of digital technologies are frequently written and communicated solely in English, potentially exacerbating existing racial and ethnic differences in cardiovascular-related health outcomes. In this work, we evaluated the language-related inclusion and exclusion criteria of digital health trials related to cardiovascular medicine registered on ClinicalTrials.gov.

**Methods:** Planned, ongoing, and completed digital health trials were identified using the ClinicalTrials.gov website interface using terms related to digital health (see https://cardio-lang-inclusion.streamlit.app/ for detailed methods and data visualization). Only digital trials testing an intervention, conducted within the United States, and studying adults were included in analyses. The identified digital health trials were further subset to select for those related to cardiovascular medicine (i.e., prevention, weight loss and cardiometabolic disease, coronary disease, vascular disease, and arrhythmias). Natural language processing was leveraged to process the 1192 digital health trials’ inclusion and exclusion criteria.

**Results:** There were a total of 1192 digital health trials related to cardiovascular medicine were identified. Over a third (470 (39.4%)) listed English language fluency as an inclusion criteria, while 196 (16.4%) excluded non-English language fluent participants. In total, 611 (51.3%) of the analyzed digital trials in cardiovascular medicine excluded participation of non-English language fluent patients (see **Figure 1**). Manual chart review of 50 random digital health trials in cardiovascular medicine demonstrated that 23 (46%) required English language fluency as an inclusion criteria and 2 (4%) excluded non-English language fluent patients from participation. These findings align with the broader automated natural language processing analysis.

**Conclusions:** We found that there was frequent exclusion of non-English language fluent patients from digital health trials in cardiovascular medicine. This may limit the generalizability of digital health trials, which is critical as we transition to decentralized clinical practice using computers and smartphones and require an accompanying rigorous evidence base.

*What is the clinical question being addressed?:* Trials in digital health use text curated by investigators. However, with the large numbers of non-English-fluent individuals in the US, these trials may exclude minority populations. Hence, we sought to investigate whether digital health trials required English fluency for participation.

*What is the main finding?:* 611/1192 (51.3%) digital health trials required English language fluency for participation. This limits the generalizability of current digital health trials. As we move toward more decentralized care, we will need a broad evidence base that includes non-English-fluent individuals.

---

Integration of digital technologies, intended to decentralize care from the clinic and/or hospital setting into patient’s homes and communities, has the potential to revolutionize clinical practice of cardiovascular medicine^1^. However, clinical trials of digital technologies are frequently written and communicated solely in English^2^, in a time when >67 million residents of the United States (US) speak a language other than English at home^3^. This is particularly deleterious for Hispanic/Latinx individuals, among whom >15.9 million indicated limited ability to understand spoken or written English^3^. Combined with the higher rates of cardiovascular risk factors in the Hispanic/Latinx population, potential exclusion of non-English language fluent participants can potentially exacerbate existing differences in cardiovascular-related health outcomes^4^.

This work was reviewed by the Stanford Institutional Review Board and deemed exempt due to retrospective analysis of public data. Here, we evaluated the inclusion and exclusion criteria of digital health trials related to cardiovascular medicine registered on ClinicalTrials.gov, to extrapolate participation of non-English language proficient individuals. Planned, ongoing, and completed digital health trials were identified using the ClinicalTrials.gov website interface using terms related to digital health, including “digital therapy”, “smartphone”, and “mobile app” (see https://cardio-lang-inclusion.streamlit.app/) for detailed methods and data visualization). Only digital trials testing an intervention, conducted within the United States, and studying adults were included in analyses. The identified digital health trials were further subset using ClinicalTrials.gov to select for those related to cardiovascular medicine via use of terms such as, “metabolic syndrome”, “diabetes”, “hypertension”, “hyperlipidemia”, “heart disease”, “arrhythmias”, and “physical activity”. Based on the search criteria, 1192 digital health trials related to cardiovascular medicine were identified. Of these, 629 were were focused on cardiometabolic disease, diabetes, and/or weight-related disorders (52.8%), 354 focused on existing cardiovascular disease including coronary, peripheral, and cerebrovascular arterial diseases and/or arrhythmias (29.7%), and 209 were focused on lifestyle and/or behavioral factor interventions (e.g., physical activity, 17.5%).

Natural language processing was leveraged to process the 1192 digital health trials’ inclusion and exclusion criteria. Over a third (470 (39.4%)) listed English language fluency as an inclusion criteria, while 196 (16.4%) excluded non-English language fluent participants. In total, 611 (51.3%) of the analyzed digital trials in cardiovascular medicine excluded participation of non-English language fluent patients (see **Figure 1**). These results did not significantly differ by disease category (e.g., cardiometabolic disease vs cardiovascular disease vs lifestyle/behavioral factors) nor by type of digital intervention (e.g., smartphone-based study vs. telemedicine vs. artificial intelligence based study). Manual chart review of 50 random digital health trials in cardiovascular medicine demonstrated that 23 (46%) required English language fluency as an inclusion criteria and 2 (4%) excluded non-English language fluent patients from participation. These findings align with the broader automated natural language processing analysis.

**Figure 1.**
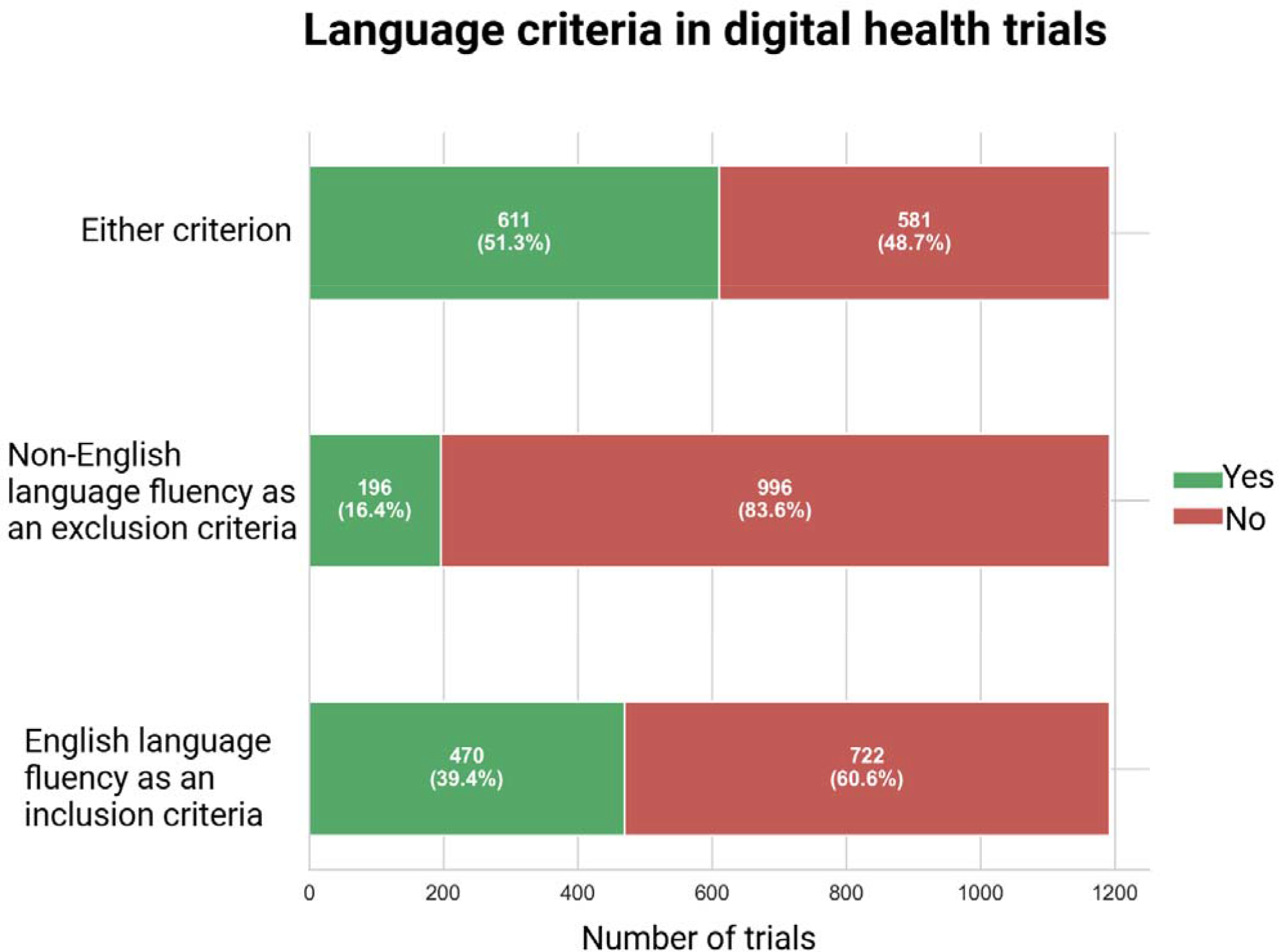
Use of English language fluency as an inclusion or exclusion criteria in clinical trials of digital health interventions (N=1196). A total of 611 (51.3%) of digital health trials used English language fluency as either an inclusion or exclusion criteria (i.e., excluded non-English language fluent patients from participation). 196 (16.4%) excluded non-English language fluent patients. 470 (39.4%) required English language fluency as an inclusion criteria for participation. Created in BioRender. Kim, D. (2025) https://BioRender.com/o46f078

We performed a separate manual review of all digital health trials that had their study protocols uploaded to ClinicalTrials.gov, but did not mention English language fluency as an inclusion criteria or exclusion criteria (N=38). On review of the study protocols, 4 (10.5%) trials required English language fluency as an inclusion criteria and 2 (5.3%) excluded non-English language fluent patients. These findings suggest that some trials may have not accurately reported their language requirements on ClinicalTrials.gov, leading to a potential underestimation of language restrictions in the natural language processing analysis.

While we note that 120 (10.0%, 120/1196) of digital trials offered translation services (with 95% of non-English support for digital trials being in Spanish), future trials of digital interventions should specifically focus on Spanish language support to increase generalizability of findings to the broader US population. This is of importance as digital interventions are increasingly adopted in the clinical realm, as a rigorous evidence base in a variety of diseases and conditions is currently needed. Moreover, continued exclusion of non-English language fluent patients from digital health trials may exacerbate existing differences in cardiovascular outcomes, particularly in the Hispanic/Latinx population that have greater existing prevalence of cardiovascular risk factors^4^.

Our analysis has several limitations. First, our analyses were based on data provided by investigators on ClinicalTrials.gov, which may be incomplete. Moreover, our search may have missed relevant digital trials that did not have appropriate search terms inputted by investigators. Second, demographics (e.g., race/ethnicity, gender, level of education, etc.) of the study population for each digital health trial could not be extracted for the present analysis based on available data on ClinicalTrials.gov. Future analysis of such data would be valuable in providing further evidence on potential differences between studied populations and the broader United States population.

In summary, we found frequent exclusion of non-English language fluent patients from digital health trials in cardiovascular medicine. This may limit the generalizability of digital health trials, which is critical as we transition to decentralized clinical practice using computers and smartphones and require an accompanying rigorous evidence base. However, with future investment in multi-lingual support, this represents a potentially solvable problem (particularly with recent advances in technology), and moreover, a solution that could potentially close the gap in health-related outcomes in cardiovascular medicine^5^.

## Data Availability

All data and code are available at a third party website (see separately).

https://cardio-lang-inclusion.streamlit.app/

