## Supplemental Methods for "Speaking the Language of Inclusion: Examining English Languages Requirements in Cardiovascular Digital Health Trials"

### **Data Acquisition and Initial Processing**

An initial dataset comprising 1,986 clinical trials, identified by their National Clinical Trial (NCT) numbers, was obtained from "clinicaltrials.gov" based on the following query (metabolic syndrome OR obesity OR overweight OR weight OR diet OR nutrition OR prediabetes OR pre-diabetes OR diabetes OR prehypertension OR hypertension or dyslipidemia OR hyperlipidemia OR heart disease OR coronary artery disease OR atherosclerotic vascular disease OR coronary atherosclerosis OR atherosclerosis, coronary OR atherosclerosis OR atheroscleroses OR atheromatosis OR cardiovascular OR peripheral artery disease OR physical activity OR exercise OR sedentary OR healthy OR lifestyle factors OR heart failure OR atrial fibrillation OR atrial flutter OR arrhythmia | digital therapeutic OR digital therapy OR digital therapies OR mobile health OR smartphone OR smart phone OR digital intervention OR mobile platform OR mobile app OR mobile device OR study app OR digital treatment OR android OR app OR app. OR app, OR digital tablet OR iOS OR iPhone OR smartwatch OR smartwatch OR virtual reality OR video game OR digital health OR mobile video OR digital platform OR software intervention OR software treatment | In United States | Adult (18 - 64), Older adult (65+) | Accepts healthy volunteers | Interventional studies). An automated procedure using natural language processing was employed to extract the participant inclusion and exclusion criteria text for each trial record from their web entry. This extraction process was subject to occasional errors, leading to incomplete criteria data for a subset of trials. Following this initial data collection and extraction, a manual curation step was performed, resulting in the removal of 498 trials that studied a pediatric population and those whose primary subject area was in either cancer or neurologic disorders. This yielded an intermediate working dataset of 1,488 clinical trial records.

### **Language Criteria Extraction and Refinement**

The extracted inclusion and exclusion criteria texts for the 1,488 trials were processed using natural language processing for each trial record from their web entry on ClinicalTrials.gov. This extraction process was subject to occasional errors, leading to incomplete criteria data for a subset of trials. An initial analysis of this automated extraction output was performed to identify trials where criteria extraction was incomplete (i.e., both inclusion and exclusion fields were blank, or only one was populated). For these identified trials, the complete inclusion and exclusion criteria were manually retrieved from ClinicaTrials.gov and integrated into the dataset. Subsequently, the language criteria extraction process was re-executed on the fully populated dataset of 1,488 trials to ensure consistent analysis.

### **Trial Categorization and Feature Identification**

Each of the 1,488 trials was then subjected to automated categorization. This analysis utilized the structured 'Conditions' field and the free-text 'Brief Description' associated with each trial record. Trials were assigned to one of several predefined main categories: Metabolic & Weight-Related Disorders, Cardiovascular Diseases, Cancers, Lifestyle & Behavioral Factors, Neurocognitive & Mental Health, or Other / Special Populations. Trials categorized under Cardiovascular Diseases were further sub-categorized into specific conditions (hypertension, heart failure, atrial fibrillation, stroke, other heart disease) using the same NLP-based approach applied to the trial descriptions.

Concurrently, this analysis assessed the trial descriptions to identify studies employing digital health technologies (defined by the mention of smartphones, wearables, or tablets).

### **Final Cohort Selection**

Based on the primary research focus, a final selection filter was applied to the categorized dataset. Only trials assigned to the main categories of "Metabolic & Weight-Related Disorders," "Cardiovascular Diseases," or "Lifestyle & Behavioral Factors" were retained for the final analysis. This step excluded 296 trials, resulting in a final analytic cohort of 1,192 clinical trials (see **Supplemental Figure 1**).

**Supplemental Figure 1. Flowchart of trial identification and filtering for analysis.**

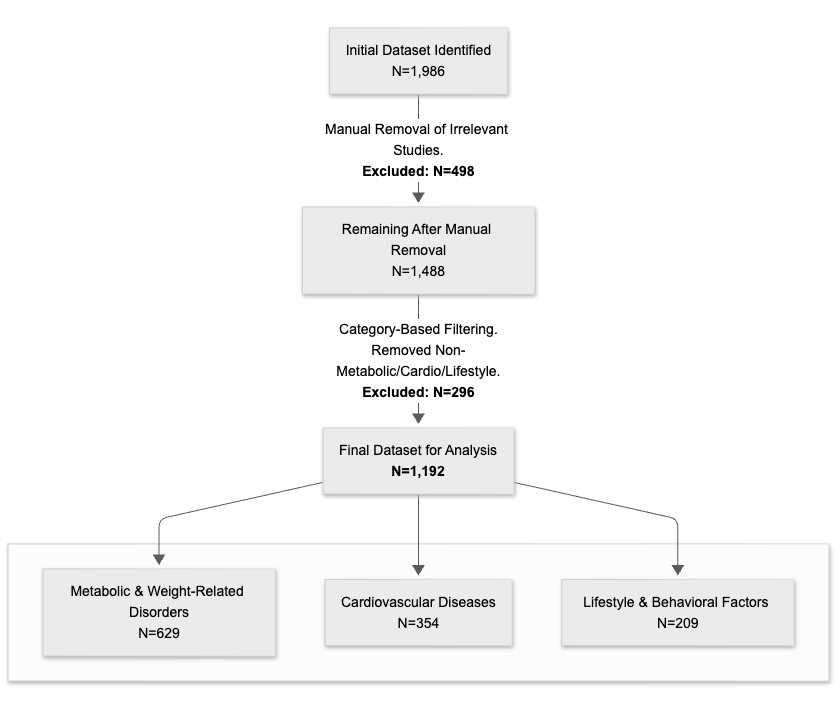
